# Convolutional Neural Network Model to Detect COVID-19 Patients Utilizing Chest X-ray Images

**DOI:** 10.1101/2020.06.07.20124594

**Authors:** Md. Shahriare Satu, Khair Ahammed, Mohammad Zoynul Abedin, Md. Auhidur Rahman, Sheikh Mohammed Shariful Islam, AKM Azad, Salem A. Alyami, Mohammad Ali Moni

## Abstract

This study aims to propose a deep learning model to detect COVID-19 positive cases more precisely utilizing chest X-ray images. We have collected and merged all the publicly available chest X-ray datasets of COVID-19 infected patients from Kaggle and Github, and pre-processed it using random sampling approach. Then, we proposed and applied an enhanced convolutional neural network (CNN) model to this dataset and obtained a 94.03% accuracy, 95.52% AUC and 94.03% f-measure for detecting COVID-19 positive patients. We have also performed a comparative performance between our proposed CNN model with several state-of-the-art machine learning classifiers including support vector machine, random forest, k-nearest neighbor, logistic regression, gaussian naïve bayes, bernoulli naïve bayes, decision tree, Xgboost, multilayer perceptron, nearest centroid and perceptron as well as deep learning and pre-trained models such as deep neural network, residual neural network, visual geometry group network 16, and inception network V3 were employed, where our model yielded outperforming results compared to all other models. While evaluating the performance of our models, we have emphasized on specificity along with accuracy to identify non-COVID-19 individuals more accurately, which may potentially facilitate the early detection of COVID-19 patients for their preliminary screening, especially in under-resourced health infrastructure with insufficient PCR testing systems and testing facilities. Moreover, this model could also be applicable to the cases of other lung infections.

## 1. Introduction

Novel coronavirus disease (COVID-19) is an ongoing pandemic caused by severe acute respiratory syndrome coronavirus 2 (SARS-CoV-2) (Stoecklin et al., 2020). The first case of COVID-19 was believed to be detected at Wuhan, China in December 2019 and it had been spread rapidly throughout the world (Holshue et al., 2020). In has been reported that viruses from the *Coronaviridae* family were discovered in 1960s from the nasal pits of patients (Lippi & Plebani, 2020). It is a massive infectious group that enveloped ribonucleic acid (RNA) viruses and generated different types of respiratory, hepatic and neurological diseases among humans and other mammals (Zhu et al., 2020; Guan et al., 2020). It contains a large family of viruses where some of them induced community transmissions, such as the middle east respiratory syndrome (MERS-CoV) and severe acute respiratory syndrome (SARS-CoV). The SARS-CoV, MERS-CoV and SARS-CoV-2 were reported to be originated from bats (Lu et al., 2020), but, however, SARS-CoV-2 has been found to have phylogenetic similarity with SARS-CoV (Zhu et al., 2020). It causes COVID-19 that makes the third coronavirus emergent condition after the past two decades, preceded by the SARS-CoV and MERS-CoV outbreak in 2002 and 2012, respectively. The World Health Organization (WHO) has announced this situation as a public health emergency of international concern on 30^*th*^ January and declared the situation as a *pandemic* on 11^*th*^ March, 2020 (Dutta et al., 2020). Moreover, WHO issued public health advises to maintain different precautions like keep social distancing, wash hand with soap and sanitizer, avoid touching nose, mouth, and eye etc. However, most of the affected countries underwent a completed locked-down to prevent the local transmission of COVID-19 infection in their regions. However, SARS-CoV-2 infected patients were commonly identified primarily with some common symptoms such as fever, cough, fatigue, loss of appetite, muscle pain etc. Hence, they were needed to identify, isolate and ensure the treatment policy at early stages. There are existing two types of procedures, namely i) molecular diagnostic tests, and ii) serologic tests (Organization et al., 2020). The reverse transcription polymerase chain reaction (RT-PCR) test is a molecular diagnostic test, which is currently considered as the gold standard (Butt et al., 2020) that detects the viral RNA of SARS-CoV-2 from sputum or nasopharyngeal swab. Nevertheless, it is relatively associated with true positive rate and required specific equipment (Butt et al., 2020). Another technique is currently under development that are explored virus proteins to identify COVID-19 called viral antigen detection. When a patient has been recovered and needed to test again, the molecular tests cannot detect this disease for longer periods as the growth of the antibody may be shown in its reaction to the host. Serologic test is another primary tool to verify antibodies in blood and diagnose patients. Due to lack of analysis and skilled human resources, these procedures are time consuming and sometimes unavailable for many people especially in low and middle income countries, which, therefore, demands an alternate but cheaper solution for early diagnostics of COVID-19 inflections fast as possible.

Recently, medical images such as chest X-ray and computed tomography (CT) scan images have been used to determine COVID-19 positive cases (Ng et al., 2020; Liu et al., 2020). But, CT scan imaging is a costly procedure and not available in every hospital or medical centers. Alternatively, chest X-ray scanning machines are found in almost all the nearest clinic, medical lab or hospitals rather than biomolecular laboratory test. So, it is cheaper, faster and widely-used way to generate 2D images of the patients ( Kroft et al., 2019), that can potentially be used for COVID-19 patients as well. Moreover, radiologists have used these images to explore its pathology and detect relevant diseases. Most of the existing works were implemented machine learning algorithms into medical images for detecting COVID-19 patients and focused on how classifiers were adopted to identify positive cases, but failed to detect false negative cases that cause more community transmission of COVID-19, requiring more stringent attention on the specificity measure of those model predictions. In this study, we proposed a convolutional neural network (CNN) to investigate chest X-ray images and identify COVID-19 patients in early stage more precisely with higher specificity, that may aid public health systems to reduce the local community transmission rate.

This paper is organized as follows: Section 2 provides some related works about chest X-ray image analysis. Then, section 3 describes about working dataset and step-by-step procedure of how we can analyze it using machine learning (ML) and deep learning (DL) models. Section 4 shows the experimental outcomes and Section 5 explains the performance of this work. Finally, Section 6 concludes this work by providing some future research directions.

## 2. Literature Review

Several studies have reported the use of medical images of COVID-19 infection for further investigation using various machine and deep learning methods. Wang et al. (2020) generated a large benchmark dataset with 13,975 chest X-ray images called COVIDx and investigated them using deep learning model that showed 93.30% accuracy. Abbas et al. (2020) proposed DeTraC deep CNN model that gave solution by transferring knowledge from generic object recognition to domain-specific tasks. Their algorithm showed 95.55% accuracy (specificity of 91.87%, and a precision of 93.36%). Apostolopoulos & Mpesiana (2020) implemented transfer learning using CNNs into a small medical image dataset (1427 X-ray images), which provided highest 96.78% accuracy, 98.66% sensitivity, and 96.46% specificity respectively. Karar et al. (2020) proposed a deep CNN architecture called COVIDX-Net that investigated 50 chest X-ray images with 25 COVID-19 cases and provided 90% accuracy and 91% F-score. Minaee et al. (2020) proposed a deep learning framework based on 5000 images named COVID-Xray-5k where they applied ResNet18, ResNet50, SqueezeNet and Densenet-121 into them and produced sensitivity 97.5% and specificity 90% on average. Heidari et al. (2020) used transfer learning based VGG16 model into chest x-ray images which showed 94.5% accuracy, 98.4% sensitivity and 98% specificity. Again, Khan et al. (2020) represented a deep neural network based on Xception named CoroNet that provided 89.6% accuracy for four class and 95% accuracy for three class images. Chandra et al. (2021) used two-phase classification approach that extracted majority vote based ensemble classifier and showed 91.03% accuracy to detect COVID-19 from pneumonia. Also, Ismael & Şengür (2021) used several deep learning approaches, namely deep feature extraction with SVM, fine tuning pre-trained CNN, and end-to-end trained CNN model, which classified COVID-19 and normal chest X-ray images. Karthik et al. (2020) proposed a customized CNN with distinctive filter learning module that shows 97.94% accuracy and 96.90% F1-score for predicting four classes respectively. Ohata et al. (2021) proposed an automatic detection model where MobileNet with the SVM (linear kernel) provides 98.5% accuracy and an F1-score and DenseNet201 with MLP shows 95.6% accuracy and an F1-score for COVID-19 infection based on chest X-ray images. Moura et al. (2020) investigated 1616 chest X-ray images using DenseNet161 where it shows 79.89% accuracy to classify normal, pathological and COVID-19 patients. Duran-Lopez et al. (2020) represented a custom CNN based model named COVID-XNet that shows 94.43% average accuracy, 98.8% AUC, 96.33% sensitivity, and 93.76% specificity respectively. Shorfuzzaman & Hossain (2020) provided a siamese neural network called MetaCOVID to integrate contrastive learning with a fine-tuned pre-trained ConvNet encoder and capture unbiased feature representations using 10-shot learning scores and compared among the meta learning algorithm with InceptionV3, Xception, Inception, ResNetV2, and VGG16. Shankar & Perumal (2020) proposed fusion model hand-crafted with deep learning features (FM-HCF-DLF) that used multi-layer perceptron (MLP) and InceptionV3 where MLP generated 94.08% accuracy.

## 3. Materials and Methods

The working methodology has been used to detect COVID-19 patients from the publicly available datasets. Figure 1 illustrates these steps that can be split into different sections such as data collection, pre-processing, classification and evaluation respectively. This approach is described briefly as follows:

**Figure 1:**
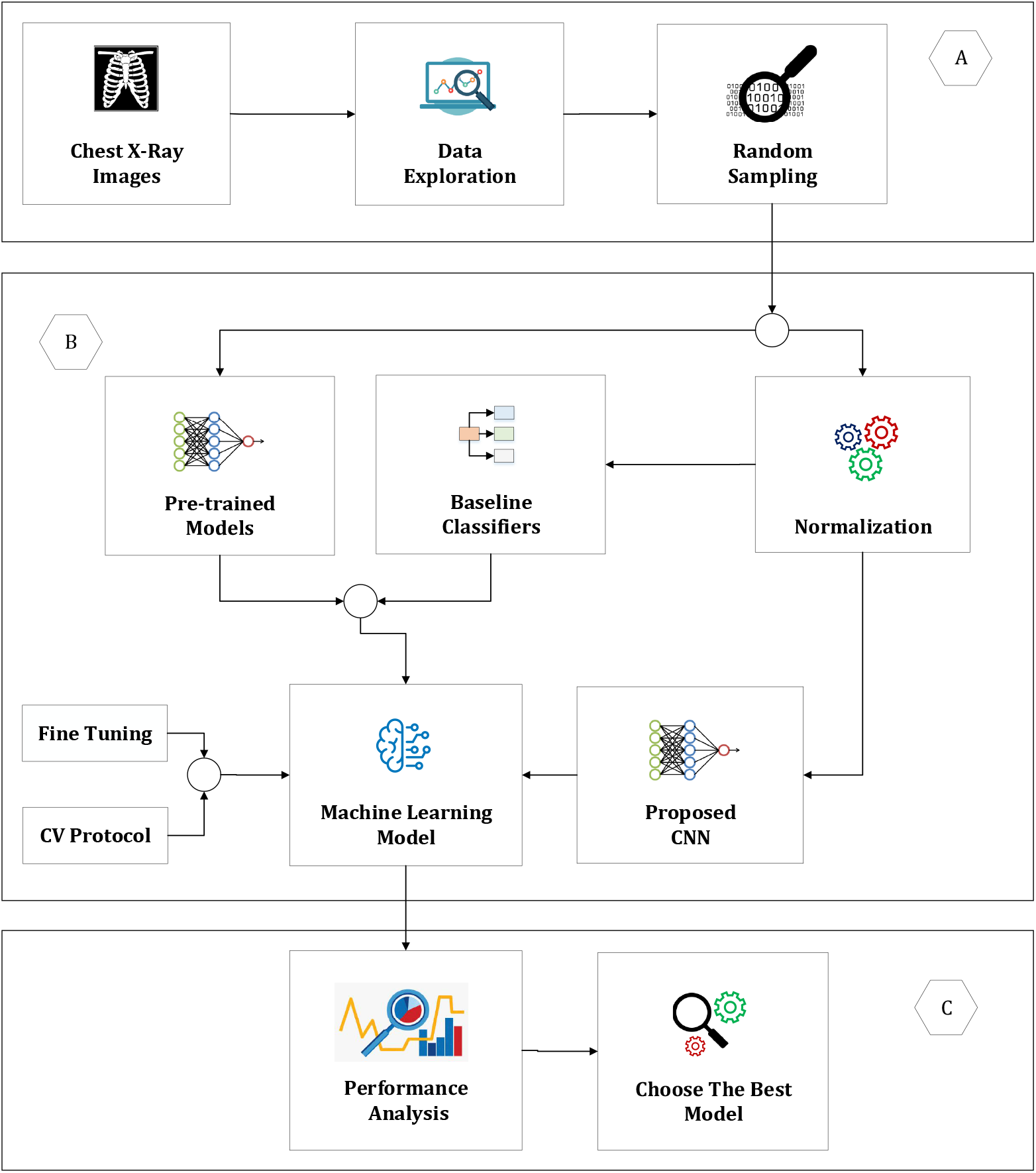
A schematic diagram of our overall workflow, where (A) chest X-Ray data has been prepossessed (B) Build a machine learning model where proposed CNN has been implemented into input images along with baseline and pre-trained models (C) Analyze the performance of proposed CNN with baseline and pre-trained models and identify the best model

### 3.1. Data Collection

The primary chest X-ray images have been obtained from the COVID-19 Radiography Database (Chowdhury et al., 2020). It contained 1,341 normal, 1,345 viral pneumonia, and 219 COVID-19 patient’s images, which have been taken as primary dataset. However, the distribution of different types of images was not the same. To balance this dataset, we collected 66 images from Cohen et al. (2020) and added them with COVID-19 images of primary dataset. For other classes (normal and pneumonia), a random undersampling method has been used and generated balanced instances of each class. Finally, this experimental dataset had been contained 285 normal, viral pneumonia and COVID-19 images respectively. For instance, Figure 2 illustrates the three types of chest X-ray images, respectively.

**Figure 2:**
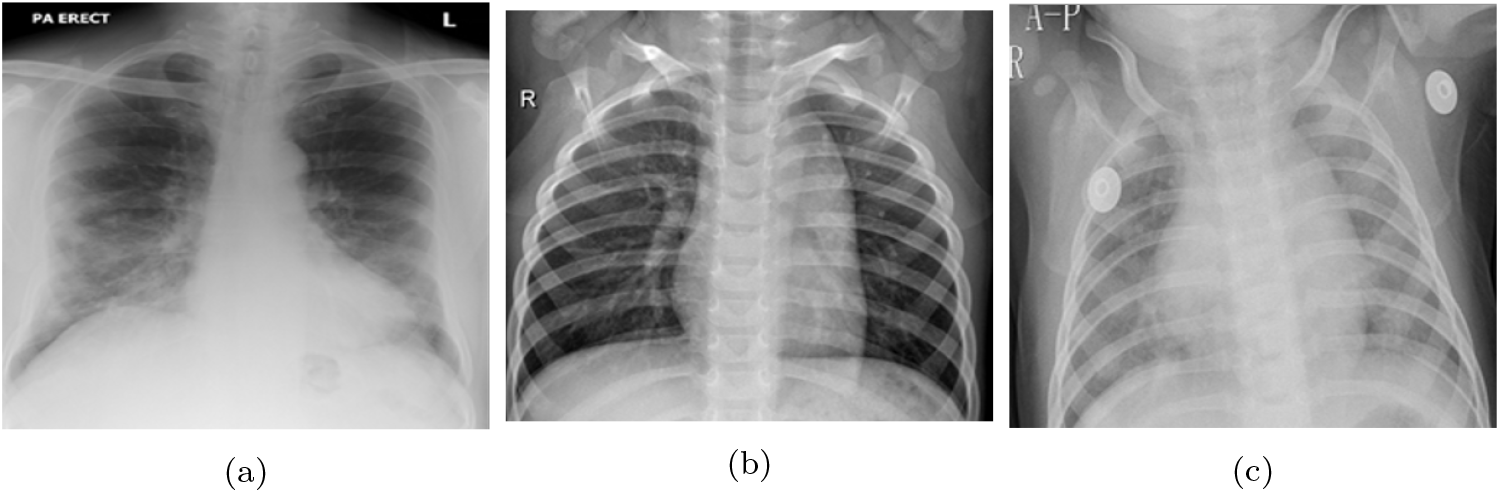
Chest X-ray images of (a) COVID-19 (b) normal and (c) viral pneumonia patients

**Figure 3:**
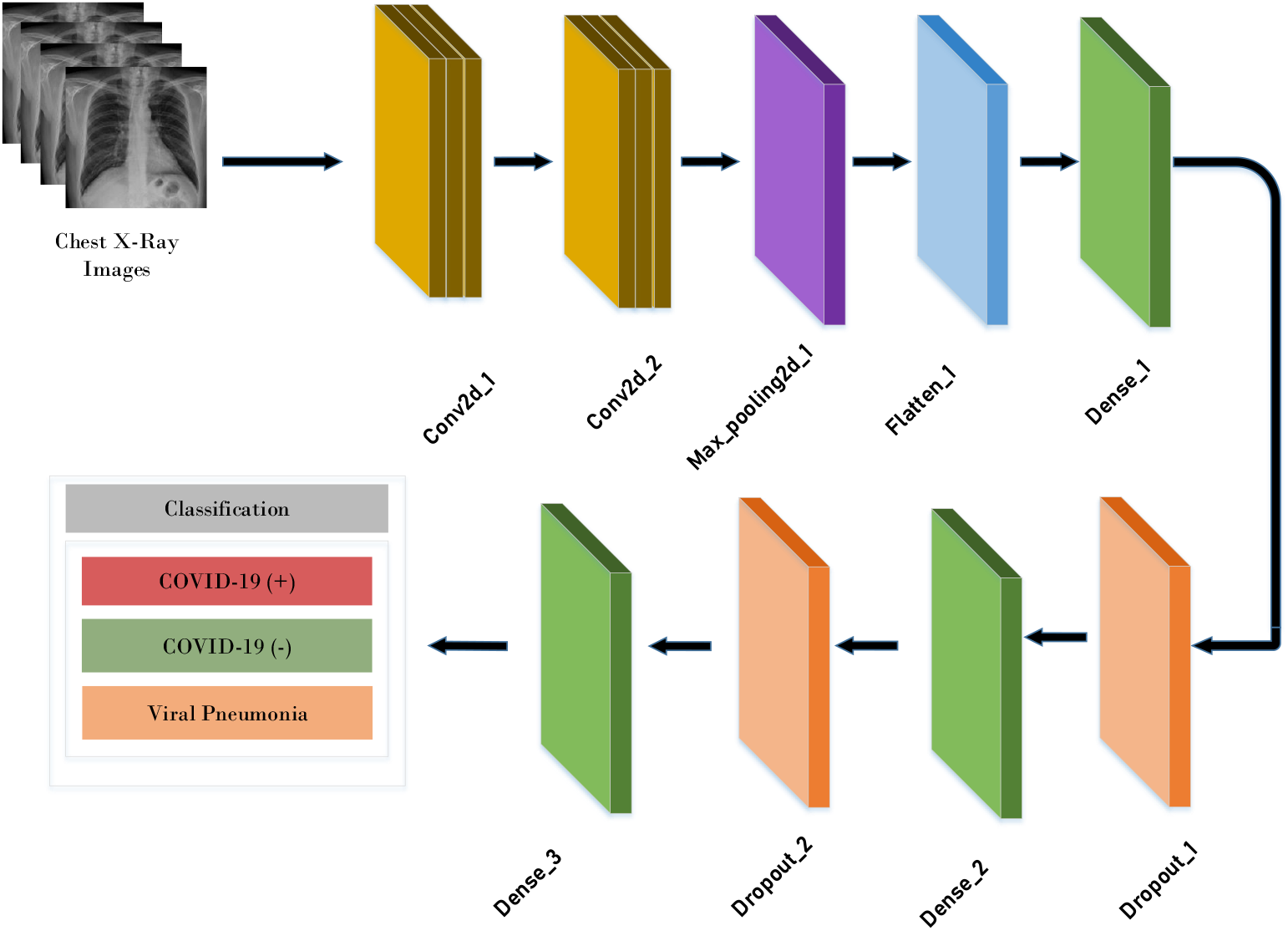
Proposed Convolution Neural Network

### 3.2. Data pre-processing

In this step, we normalized training set into grayscale images. Then, all baseline classifiers have been implemented with transformed dataset respectively. But, pre-trained CNN models such as VGG16, ResNet50, InceptionV3 cannot support grayscale images, hence we directly employed them into primary dataset.

### 3.3. Proposed Convolutional Neural Networks

Convolutional Neural Networks (CNN) is a special class of artificial neural network (ANN) that manipulates an input layer along with the sequence of hidden and output layers. It maintains a sparse connection between layers and weights that shares them with output neurons in the hidden layers. Like regular ANN, CNN contains a sequence of hidden layers, which are denoted as convolutional and polling layer. In addition, the operations of these layers are called convolutional and polling operation, respectively. Alternatively, they are stacked to lead a series of fully connected layers followed by an output layer. In many research fields including image recognition, object detection, semantic segmentation and medical image analysis, CNN models yield considerably higher performances compared to the state-of-the-arts.

#### 3.3.1. Convolutional Layer

Convolution layer is the core structure of a CNN that manipulates convolution operation (represented by *) instead of general matrix multiplication. This layer accomplishes most of the computations of CNN model. The count of filters, size of local region, stride, and padding are mentioned as *hyper-parameters* of this layer. Convolution layers extract and learn about features using these filters. Hence, it is known as the feature extraction layer. These parameters are necessary where the similar filter is traversed across the whole image for a single feature. The main objective of this layer is to identify common features of input images and map their appearance to the feature map. The convolution operation is given as:

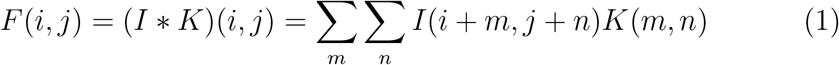

To introduce non-linearity, the output of each convolutional layer is fed to an activation function. Numerous activation functions are available but Rectified Linear Unit (ReLU) is widely used in the deep learning field. It is mathematically calculated as follows:

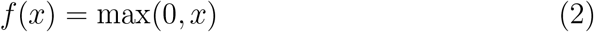

In this model, we have used fewer layers and filters, which consists of two convolutional layers and gradually increased the number of filters from 32 to 64, respectively, where an image of size 100×100 and the *pixel* values of them are either 0 or 1. In the first convolutional layer, this image is convoluted with 3 *×* 3 kernel for 32 filters and produces the feature map 100 *×* 100 *×* 32. Subsequently, this output has been forwarded to the second convolutional layer where we consider 3 *×* 3 sized kernel for 64 filters that is also convoluted with 100 *×* 100*×* 32 extracted features and produced 100 *×* 100 *×* 64 sized output feature map is produced in this layer.

#### 3.3.2. Pooling Layer

In CNN, the sequence of convolution layer is followed by an optional pooling or down sampling layer to lessen the volume of input images and number of parameters. This layer computes fast and precludes over-fitting. The most common pooling technique is called *Max Pooling*, which merely generates the highest result of the input region. Other pooling options are *average pooling* and *sum pooling*. Two hyper-parameters are essential for the pooling layer, namely filter and stride. In this model, we implement 2 *×* 2 filter into 100 *×* 100 *×* 64 sized output feature map and create 50 *×* 50 *×* 64 reduced feature map.

#### 3.3.3. Flatten layer

After implementing the pooling layer, a flatten layer has been employed to flat the entire network. It converts the entire pooled feature map matrix into a single column.

#### 3.3.4. Dense layer

Then, we have implemented three dense layers which are also known as a fully connected layer. In this layer, the input of previous layers is flattened from a matrix into a vector and forwarded it to this layer like a neural network. This layer viewed the output of past layers and decided which features are mostly matched with the individual class. Therefore, a fully connected layer can yield accurate probabilities for the different classes. The outputs are classified by using the activation function at the output layer, which in our case was the *Softmax function* to calculate the probability of particular classes defined by the following this equation:

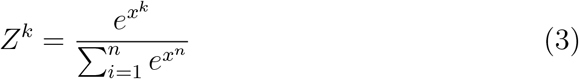

#### 3.3.5. Dropout layer

When a large *feed-forward* neural network is investigated with a small training set, it usually shows poor performance on held-out test data, and *dropout* is a useful procedure to mitigate this problem. In our model, we used dropout layer after each dense layer and to reduce over-fitting by preventing complex co-adaptations on the training data.

### 3.4. Baseline classifiers

Several machine learning classifiers have been used to perform comparative performance assessements, such as support vector machine (SVM), random forest (RF), k-nearest neighbor (k-NN), logistic regression (LR), gaussian naïve bayes (GNB), Bernoulli naïve bayes (BNB), decision tree (DT), Xgboost (XGB), multilayer perceptron (MLP), nearest centroid (NC) and perceptron. While training with pre-processed dataset, several model hyper-parameters were fine-tuned (i.e. changed and optimized) to get better predictive accuracy. These parameters of baseline classifiers are represented in Table 2.

**Table 1:**
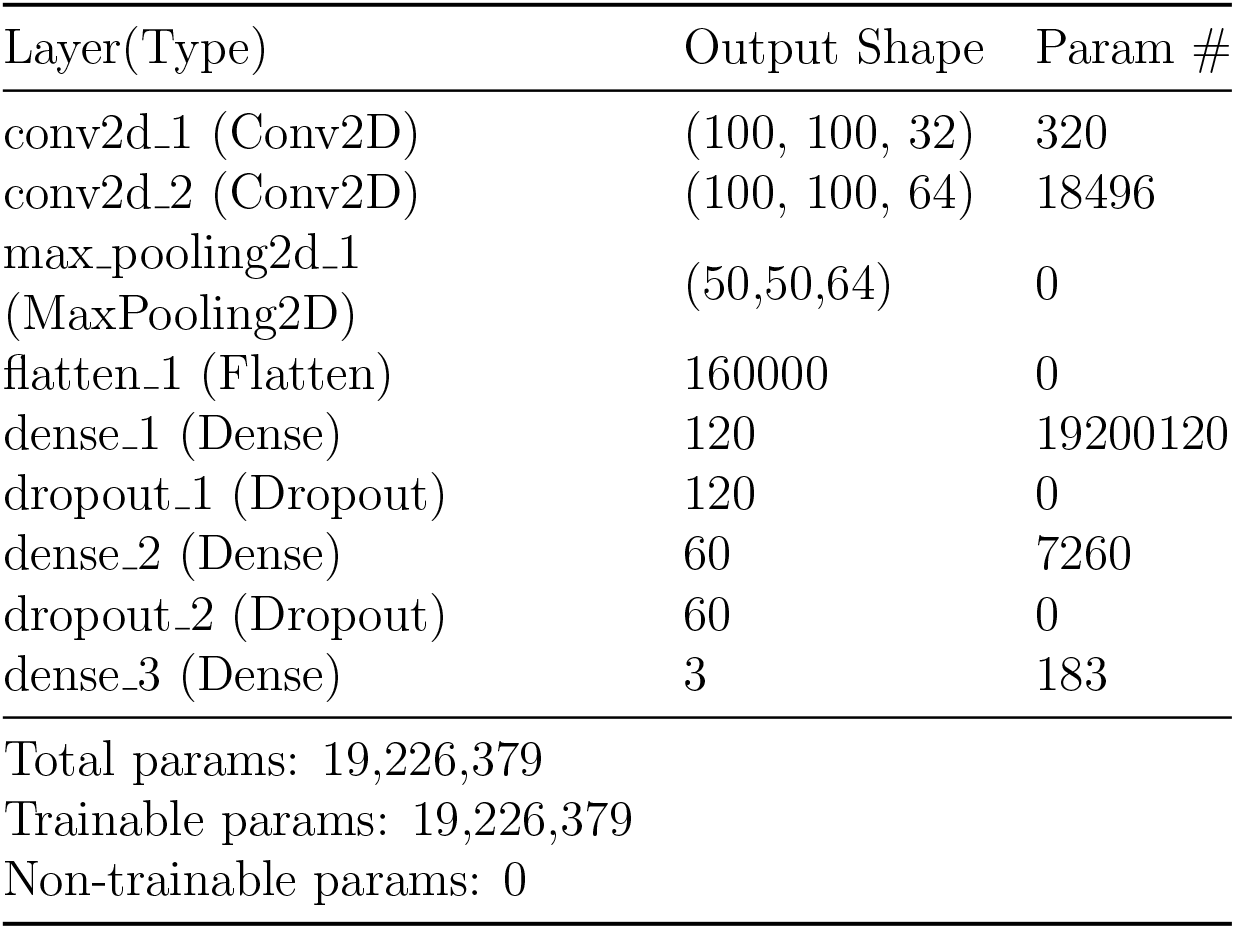
A summary of proposed 9 layers model

**Table 2:** Different model parameters of classical machine learning classifiers

| Classifier | Parameters |
| --- | --- |
| SVM | linear kernel, gamma = 0.0001 |
| KNN | k= 5, euclidean distance |
| GNB | priors = None, var_smoothing = 1e^-09 |
| BNB | alpha = 1.0, binarize = 0.0 |
| DT | gini criterion, best splitter |
| LR | liblinear solver, max_iter = 1000 |
| RT | max_depth = None, random_state = 0 |
| GB | max_features = 2, max_depth = 2,<br>random_state = 0 |
| XGB | learning_rate =0.1, max_depth = 3 |
| MLP | adam solver,<br>alpha = 1e^-5, random_state = 1 |
| NC | manhattan metric |
| Perceptron | tol = 1e^-3, random_state = 0 |

### 3.5. Pre-trained transfer learning models

Moreover, several deep learning classifiers were employed such as deep neural network (DNN) and several pre-trained CNNs like residual neural network (ResNet50), visual geometry group network 16 (VGG16), and inception network V3 (InceptionV3) for transfer learning. These models have been widely used to investigate images in various domains (Ahammed et al., 2020; Satu et al., 2020, 2019). Like general classifiers, various parameters were tuned to get more accurate result for detecting COVID-19. For DNN, we considered the batch-size as 32, number of epochs as 50, the ‘adam’ optimizer, and the learning rate as 0.0001 with *weight decay*. Again, some regularization terms have been employed for reducing overfitting in the deep learning models. When pre-trained models have been loaded, it was downloaded requiring packages to manipulate input images. Then, the flatten layer was added into these pre-trained models, which flattens the input to one dimension. Then, we implemented a dense layer with 64 neurons, the ‘*relu*’ activation function and the regularizer as 0.001, respectively. Before and after employing the dense layer, dropout layer has been used to reduce the over-fitting issues. Finally, three classes have been assigned with the ‘*softmax* ‘activation function. To compile the model, categorical crossentopy loss function and adam optimizer were taken with 0.00001 learning rate. Therefore, we used last 1 trainable layer for ResNet50 and considered the last 62 trainable layers for InceptionV3.

### 3.6. Evaluation

The performance of individual classifiers was assessed by different evaluation metrics such as *accuracy, AUC, F-measure, sensitivity*, and *specificity*, respectively. The brief description of them is given in Table 3.

**Table 3:**
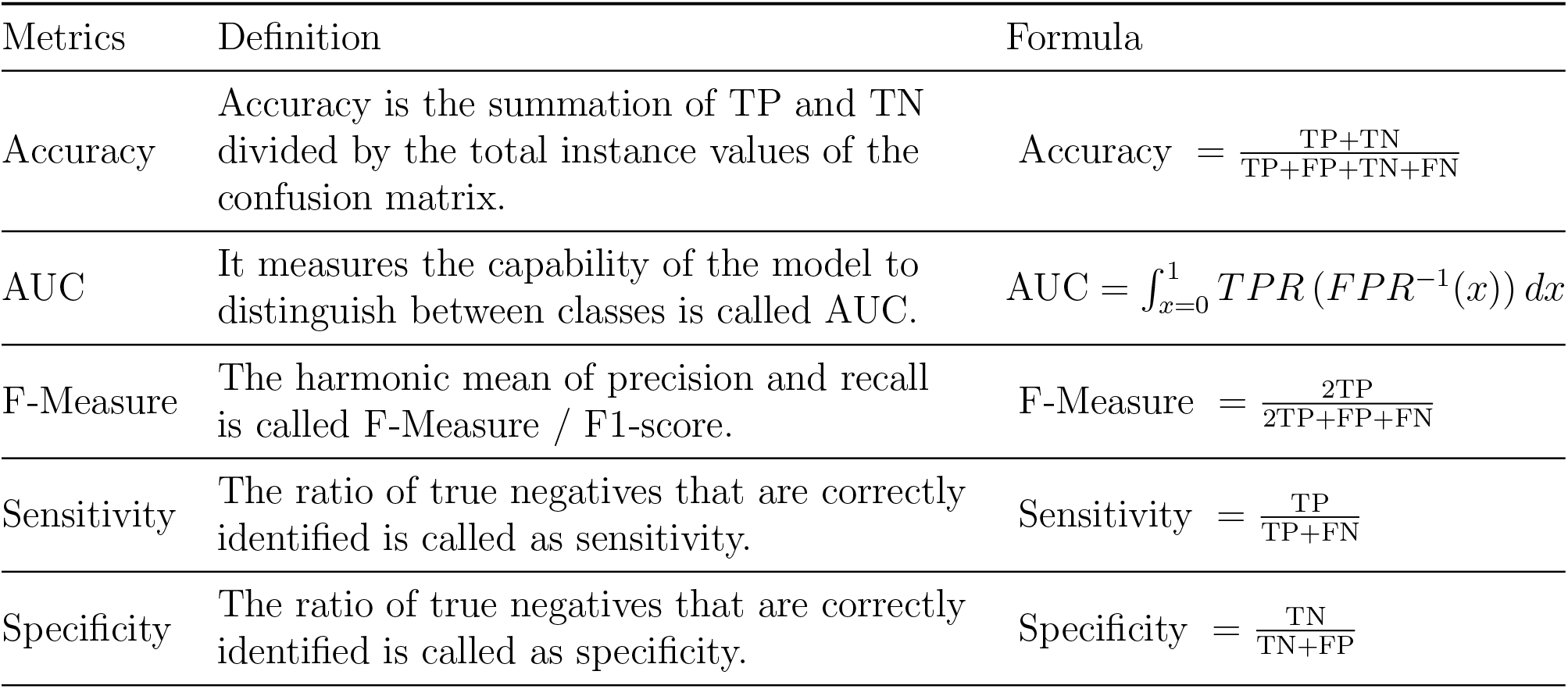
Evaluation Metrics

| Metrics | Definition | Formula |
| --- | --- | --- |
| Accuracy | Accuracy is the summation of TP and TN divided by the total instance values of the confusion matrix. | $\text{Accuracy} = \frac{\text{TP} + \text{TN}}{\text{TP} + \text{FP} + \text{TN} + \text{FN}}$ |
| AUC | It measures the capability of the model to distinguish between classes is called AUC. | $\text{AUC} = \int_{x=0}^1 \text{TPR}(\text{FPR}^{-1}(x)) dx$ |
| F-Measure | The harmonic mean of precision and recall is called F-Measure / F1-score. | $\text{F-Measure} = \frac{2\text{TP}}{2\text{TP} + \text{FP} + \text{FN}}$ |
| Sensitivity | The ratio of true negatives that are correctly identified is called as sensitivity. | $\text{Sensitivity} = \frac{\text{TP}}{\text{TP} + \text{FN}}$ |
| Specificity | The ratio of true negatives that are correctly identified is called as specificity. | $\text{Specificity} = \frac{\text{TN}}{\text{TN} + \text{FP}}$ |

## 4. Experiment Results

In this work, proposed CNN was used to investigate chest X-ray images of normal, pneumonia and COVID-19 patients. We have used various classifiers, e.g. SVM, RF, KNN, LR, GNB, BNB, XGB, MLP, NC and perceptron using *scikit* learn library in python. Again, deep learning model such as DNN and pre-trained CNN models (VGG16, MobileNet, ResNet50) were implemented using *keras* library. Therefore, these classifiers were employed 10 fold cross validation procedure using python programming language. All applications have been implemented on a laptop with Asus VivoBook S, Core i5-8250U Processor 8th generation - (8 GB/1 TB HDD/Windows 10 Home/) Intel UHD Graphics 620.

The performance of all classifiers was assessed by different evaluation metrics: accuracy, AUC, F-measure, sensitivity, and specificity respectively. The results of these classifiers are shown in Table 4. Most of the models yielded substantially good predictive performances to classify chest X-ray images, where seven of them such as Proposed CNN, XGB, LR, SVM, MLP, RF and GB show the results greater than 90%, six classifiers like perceptron, KNN, GNB, NC, DT and DNN represent their performance greater than 70% and less than 90%. Consequently, the rest of the classifiers for instances, BNB and pre-trained CNNs such as ResNet50, VGG16, InceptionV3 are provide less than 70% outcomes. The characteristics of these neural network/regression-based classifiers are more realistic to investigate COVID-19 dataset. The details of average results of different classifiers are shown in Figure 4. Among all of these classifiers, proposed CNN shows the highest accuracy (94.03%), AUROC (95.53%), F-measure (94.03%), sensitivity (94.03%), and specificity (97.01%), where it classifies 272 COVID-19 instances from 285 instances accurately (see Table 5). Then, XGB, LR, SVM, MLP, RF and GB demonstrate better results than other algorithms except for CNN. Moreover, the average performance of all classifiers is satisfactory for all evaluation metrics. We believed, due to the small number of COVID-19 cases, these deep classifiers cannot show more accurate results like CNN and others. Therefore, our proposed CNN model is found the best classifier to identify both COVID-19 positive and negative cases with high performance and can assist physician and policy maker to identify them quickly and take necessary steps.

**Table 4:**
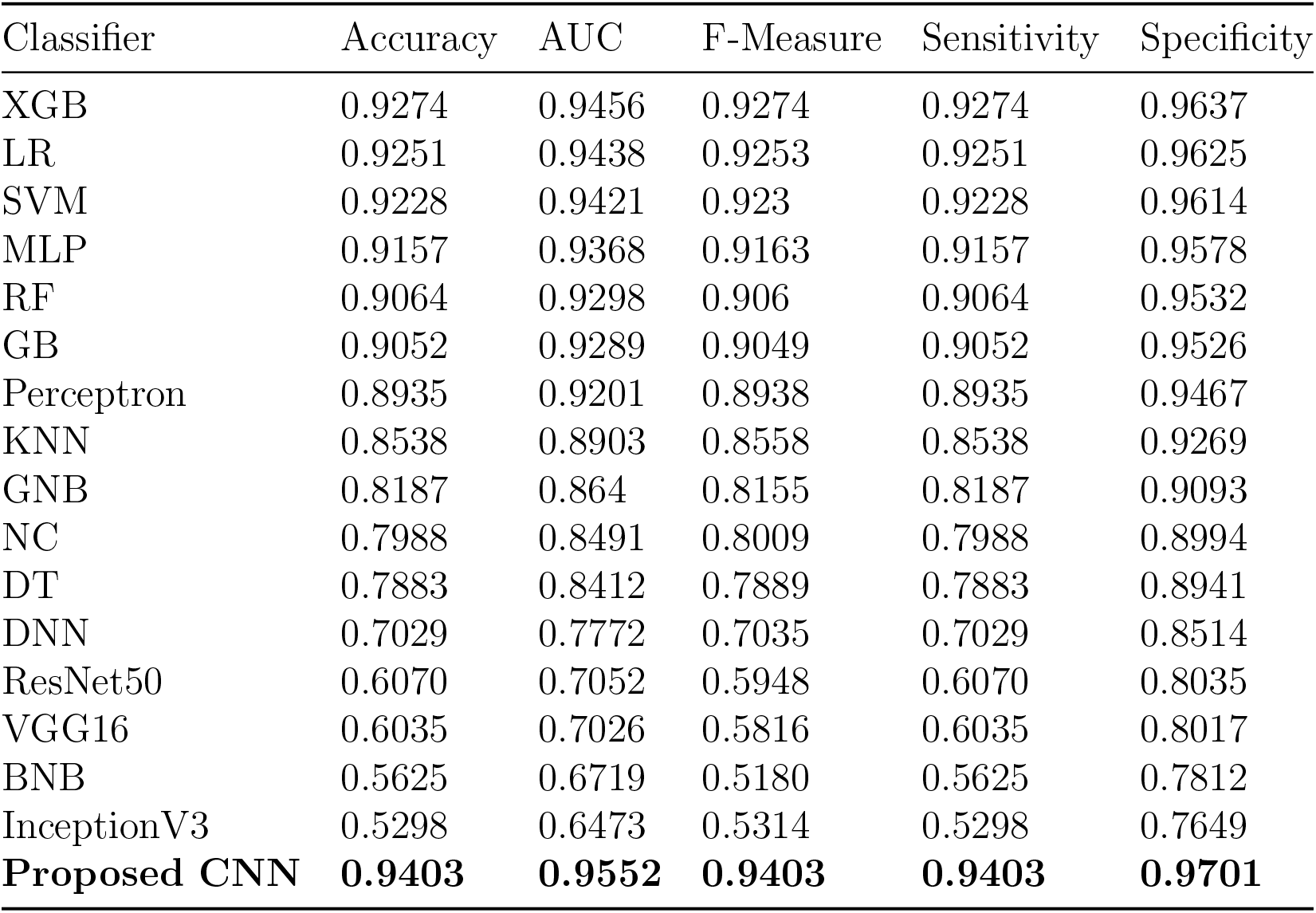
Performance Analysis of Classification Methods

**Table 5:**
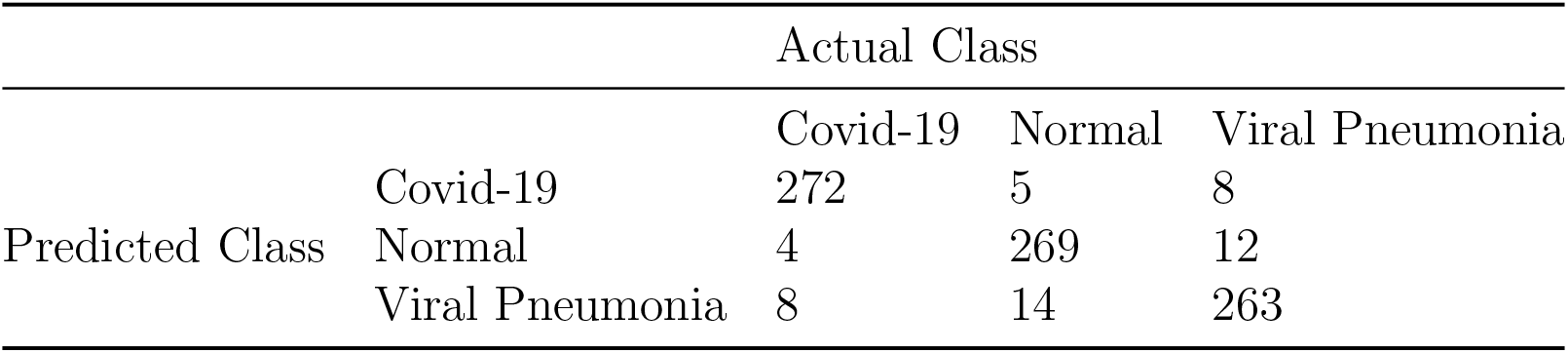
Confusion Matrix of CNN

**Figure 4:**
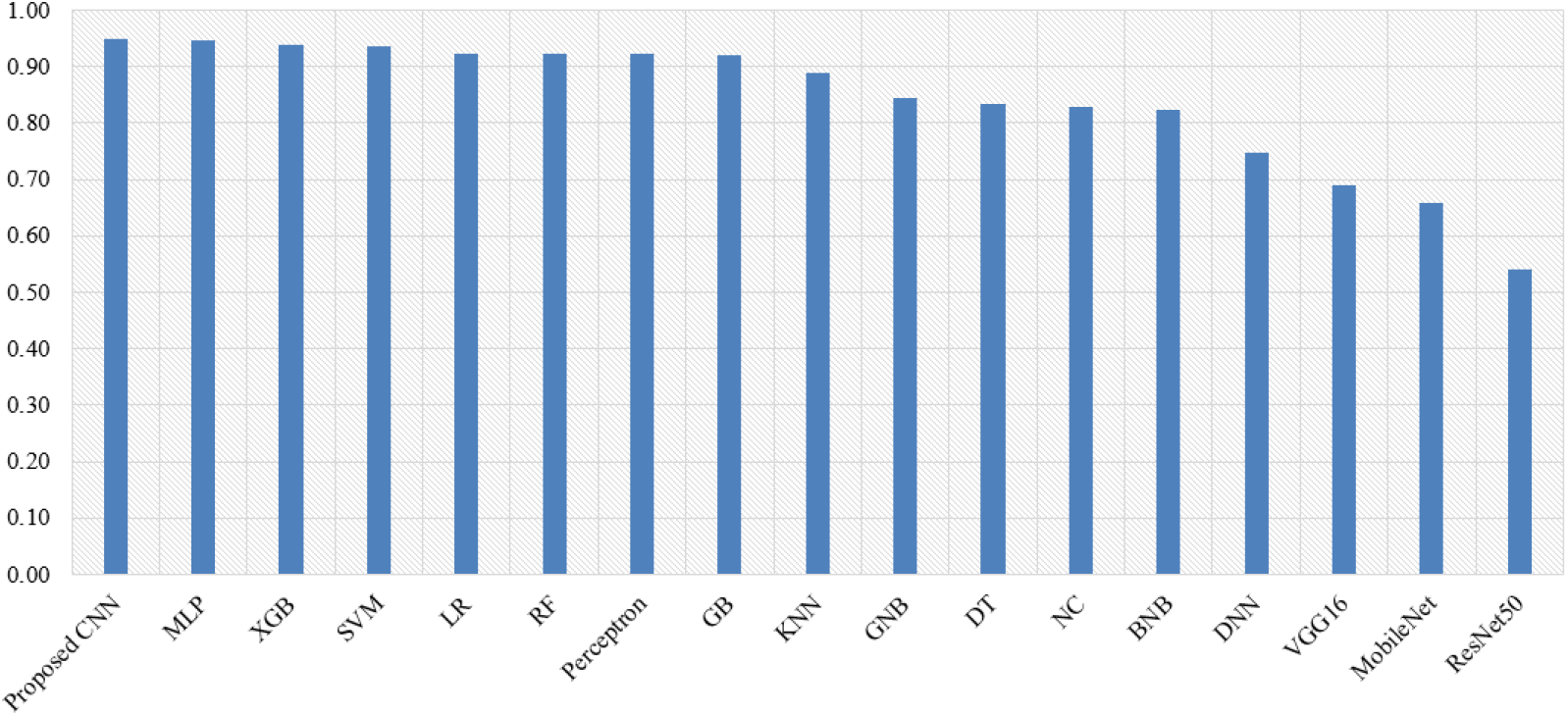
Average results of individual classifier

In the proposed CNN, some metrics like accuracy, f-measure and sensitivity are indicated how COVID-19 positive (target) cases can be determined.

Besides, they have been effective to explore and identify the target cases. This model shows 94.03% accuracy, f-measure and sensitivity and 95.52% AUC correspondingly. Besides, specificity is one of the most important terms because it shows how COVID-19 negative patients can be explored more accurately. The incremental result of specificity is denoted more appropriate identification of COVID-19 negative cases. Hence, the community transmission has been reduced by these infectious persons whose were not detected as positive cases precisely. Note, our proposed CNN has yielded the highest specificity (97.01%) among all the models evaluated in this work by outperforming them (Table 4). Thus, the proposed CNN indicate its potentiality to avoid false positive cases.

### 4.1. Comparison with Existing Works

In this work, several machine and deep learning classifiers were used to investigate chest X-ray images and detect COVID-19 positive cases rapidly. Most of the classifiers used in our work were widely implemented in earlier works with good predictive performances. Recently, many studies have been conducted related to COVID-19 chest X-ray image analysis, where some limitations were identified. Most of the works were focused on high sensitivity, i.e. how the classifiers can identify COVID-19 positive cases frequently (Karar et al., 2020; Karthik et al., 2020; Khan et al., 2020; Ismael & Şengür, 2021; Ohata et al., 2021). But, nowadays the community transmission is a great issue to prevent the spread of COVID-19 and the growth of false negative rates are accelerated, which is a great concern. So, in this work, we particularly focused on specificity (i.g. reduced false negative rates) along with other metrics. Our proposed CNN model shows better specificity (97.01%) than many existing works (Abbas et al., 2020; Apostolopoulos & Mpesiana, 2020; Minaee et al., 2020; Chandra et al., 2021; Duran-Lopez et al., 2020; Pandit & Banday, 2020). At the application level, the usage of our model may prevent COVID-19 community transmission by detecting false negative cases more accurately. In this work, we verified experimental results using various evaluation metrics like accuracy, AUC, F-measure, sensitivity, and specificity respectively. Several works have analyzed a few number of COVID-19 samples along with other cases where the experimental dataset was remained imbalanced (Karar et al., 2020; Sekeroglu & Ozsahin, 2020; Shankar & Perumal, 2020; Zebin & Rezvy, 2020). Sometimes, they were conducted with a separate dataset where the scarcity of samples was found in both of these datasets (Apostolopoulos & Mpesiana, 2020; Shankar & Perumal, 2020). In the proposed model, we integrated COVID-19 samples of related datasets in addition to balance the target classes named normal and viral pneumonia using random *under-sampling* in view of these samples. Some works have improved their results, specially, accuracy by merging non-COVID classes (e.g., class 3 to 2) (Apostolopoulos & Mpesiana, 2020). It is realized that normal and viral pneumonia are hardly associated with COVID-19. In our current study, we considered these conditions and analyze them to justify this epidemic situation. Many state-of-the-arts of CNN, such as pre-trained transfer learning models are available, which investigated chest X-ray images to classify COVID-19 (Apostolopoulos & Mpesiana, 2020; Abbas et al., 2020; Pandit & Banday, 2020). But, they were not shown better results for only 855 samples. If we will increase the number of images in another condition, it decreases the risk of false positive rate. In recent works, none justify their work with machine learning based classifier along with deep learning (Apostolopoulos & Mpesiana, 2020; Karar et al., 2020). Besides, our approach provides a solution for gathering small and a large number of chest X-images to investigate COVID-19 cases.

## 5. Discussions

Many techniques have been using the RT-PCR test and viral antigen detection techniques to identify COVID-19 cases, but none reported to claim the 100% accuracy in this case. Besides, most of this existing process are cost-heavy, time-consuming and requires specific instructions to implement them. However, sample collection is from a large population is a slow process and hence, the infection may remain undetected. Instead, chest-X-ray images are more accessible than other diagnostic measures. To tackle community transmission, in addition to better predictability, the testings should also be vigilant against false negative prediction as that undetected cases will further trigger more infections. Physicians and healthcare workers cannot take proper steps when many patients are admitted to the hospital. If these cases are detected at an early stage, they can isolate themselves from the community and give possible treatment rapidly that reduces the transmission rate of COVID-19. Therefore, we needed a suitable tool to detect COVID-19 positive and negative cases more feasible way. Our proposed CNN model can automatically detect these cases more accurately with high specificity (see Section 4.1). In this work, we focused on COVID-19 negative cases so that community transmission is unchained as strongly as possible. Besides, due to social lock-down, collection of large-scale medical images along with chest-X-ray images and investigate them using machine learning models in this pandemic situation may not be feasible. But, our proposed CNN is capable of yielding higher predictive performance even though being trained with a small number of samples compared to the existing works.

The pandemic condition getting severe day-by-day in recent times.Different sectors such as agriculture, business, finance are impeded rapidly during this period. Many people lose their jobs and do not manage new oppor-tunities due to the lock-down situation. SARS-CoV-2 transmission rate is extremely high, and hence, any undetected COVID-19 case may potentially spread out this disease throughout their community rapidly. Therefore, early detection via cost-effective tools with high predictive power is highly required to recognize cases and take proper steps as soon as possible. High specificity from our proposed CNN model could successfully reduce false negative rates by detecting subtle cases, that can significantly affect not only the public health but consequently re-induce the social and economic normality.

## 6. Conclusion and Future Work

The study proposed a CNN model, which analysed chest X-ray images of COVID-19, healthy and other viral pneumonia patients to classify and diagnosis COVID-19 patients automatically in a short period of time. Again, various machine and deep learning-based approach are used to compare the performance and the outcomes of our proposed CNN model. Hence, proposed CNN yielded the highest 94.01% accuracy and 97.01% specificity, while most of the deep learning methods did not provided good results like general classifiers. Despite taking all the measures of avoiding over-fitting, the results of our proposed CNN model performed surprisingly well with small datasets, however, it would be interesting to see its performance with larger training dataset. Hence, in future, we will collect a large number of images from various sources and analyze them to get more feasible outcomes. This approach may be helpful for clinical practices and detection of COVID-19 cases to prevent future community transmission.

## Data Availability

The link of data availability is given below:
https://github.com/shahriariit/COVID-19-Xray

## Notes

### Competing Interest Statement

The authors have declared no competing interest.

### Funding Statement

We have not got any fund to do this project

### Author Declarations

1) this material has not been published in whole or in part elsewhere; 2) the manuscript is not currently being considered for publication in another journal; 3) All authors have been personally and actively involved in substantive work leading to the manuscript, and will hold themselves jointly and individually responsible for its content.

